# Age at First Menstruation in Girls Attending Primary and Secondary Schools in Khartoum North, Sudan

**DOI:** 10.1101/2025.05.04.25326955

**Authors:** Mohamed Abdel Bagi Abdel Ghani Babiker, Mohamed Ahmed Ali ElSheikh

## Abstract

**Background:** Menarche is the stage of entering adulthood and it has a psychological and social side effects. Together with thelarche and adrenarche they make the triad of puberty in females. In Sudan, there are some published papers about age at menarche and some of its associations but the need is there for more.

**Objectives:** We aimed in this study to figure out the mean age at menarche in primary and secondary high school girls in Khartoum north city-Khartoum state, In addition, to correlation of weight, height, plus socioeconomic status with age at menarche.

**Methods:** It was a community based cross-sectional study, conducted from December 2013 to February 2014. The tool used was an interview questionnaire, to collect data from randomly selected primary and secondary high schools, in Khartoum North city. The calculated sample was 367 girls. They were interviewed about their sociodemographic data, their age at first menstruation, plus other related variables. Their weight and height were measured. Data were analyzed using IBM SPSS version 20. The mean age at menarche was calculated and a correlation with Spearman’s coefficient was done between age at menarche and the three variables weight, height and socioeconomic status.

**Results:** the girls mean age at menarche was 12.73 (SD = 0.89) years, there is a weak negative correlation with weight and socioeconomic status, and a weak positive correlation with height.

**Conclusion:** compared with previous studies in Sudan there is a tendency for a decrease in age at menarche, further studies are needed to confirm or oppose this finding.

## Introduction

Menarche is usually defined as the age of the first menstrual cycle. Furthermore, it is the stage of entering adulthood and it has a psychological and social side effects. Together with thelarche and adrenarche they make the triad of puberty in females. For puberty to start there has to be intrinsic and extrinsic factors. These factors include genetic, race, ethnic group, gender, body weight, height, nutrition, stress and exercise.^[1, 2]^ Some of these factors have been well studied in the literature and some needs further research. An example is the genetics of the age of menarche, which needs more meticulous investigation to reach applicable conclusions.^[3]^ The relationship between the above mentioned factors and age at menarche is complex, that entails the surveying of age at menarche on a regular basis to try to explore this relationship and its consequences.

In this study we aimed at determining the age at menarche of school girls in Khartoum North City, to try to increase our national knowledge about the age at menarche in Sudanese females. Few studies with scientific limitations were conducted in Sudan.

## Methods

A cross-sectional community based study where 367 primary and secondary school girls were selected randomly, and their age range was 9 to 18 years old. The sample size was calculated using Richard’s Ginger Formula. One primary and one secondary schools were chosen randomly in seven districts of Khartoum north city (one of the three big cities in Khartoum state in Sudan). The study was conducted from December 2013 to February 2014. A face-to-face interview was conducted using a pre structured questionnaire to collect data on their demography and socioeconomic status (according to their parents level of education and occupation), their age at menarche, and their age at first regular cycles. They were also asked about their parity order and class grade at school. In addition their height and weight were measured. As finding the age at menarche of these girls was the primary objective, finding its relation to the socioeconomic status and weight of the girls were one of the secondary objectives. After explanation of the research objectives, we took an informed consent from the schools headmistresses and they in turn took consent from the girl’s guardians after explaining the research idea to them. In addition, we took ascent from the girls themselves. An agreement to start the research was taken from both the primary schools and secondary schools administrations at the Khartoum state Ministry of education; in addition, their ethical committee approved the study. The data were analyzed using the IBM statistical package for social sciences version 20. The mean age, age at menarche, age at first regular cycles and mother’s age at menarche were calculated, plus Spearman correlation test to determine the relationship between the menarcheal age and weight plus the socioeconomic status of the participants girls.

## Results

All the 367 girls has been interviewed. Table 1 provides the summary statistics of the main variables of the study. It is apparent that the mean age at menarche is 12.73 ± 0.89 years, and the girls had their first regular cycles at age of 13.06 ± 1.10 years. 83 girls did not recall these development stages. In addition, figure 1 and 2 presents the distribution of the previous two variables within the participants. Moreover, an 80% of the candidates are of low socioeconomic status (SES); the other 20% are of high SES. 83.4% of them were born in urban areas while 16.6% were born in rural areas. Interestingly, 36.24% of the sampled girls were in the second class of the secondary high school, while 11.44% were in the eighth class of the primary school. In addition to that, most of the girls were born first or second in order in their families (24.52% and 22.34% respectively).

**Table 1.**
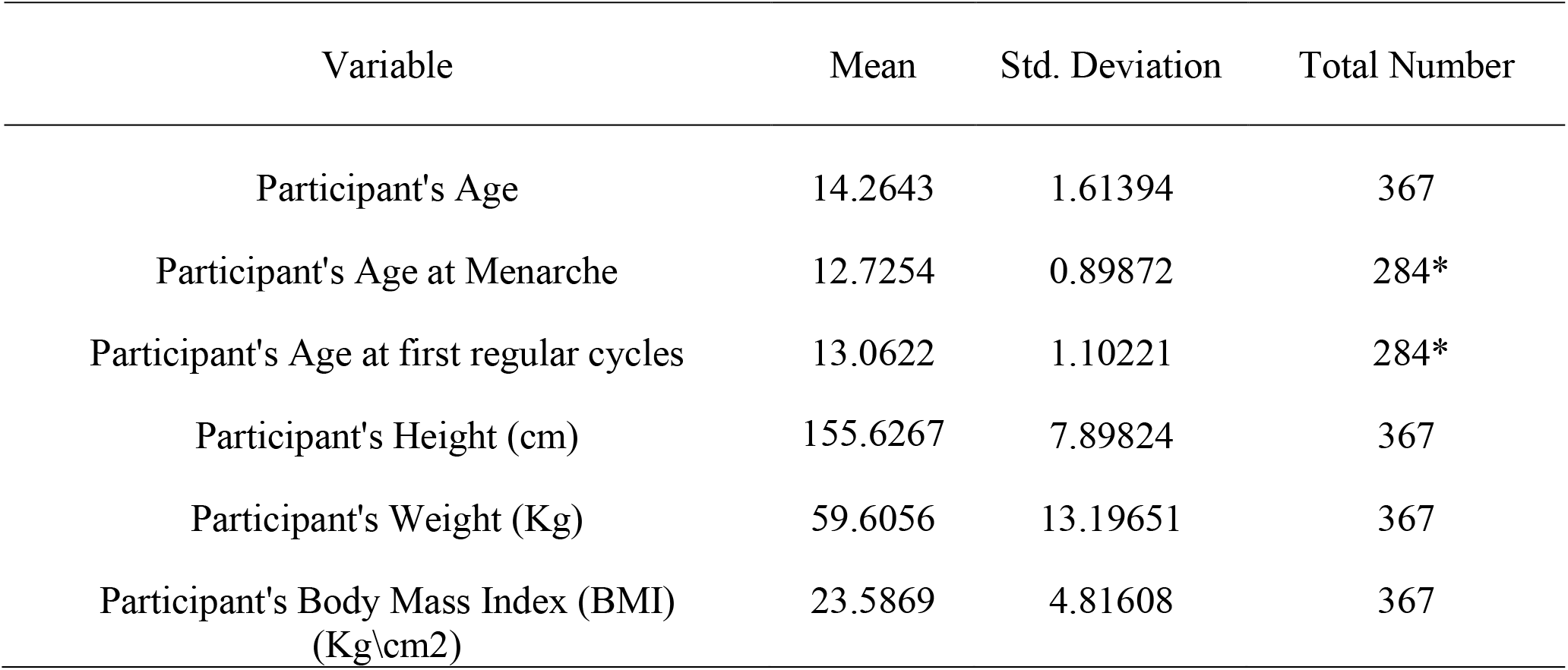
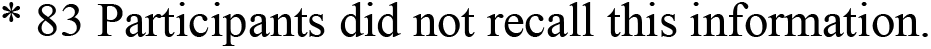
Mean and standard deviation for the main variables of the study.

| Variable | Mean | Std. Deviation | Total Number |
| --- | --- | --- | --- |
| Participant's Age | 14.2643 | 1.61394 | 367 |
| Participant's Age at Menarche | 12.7254 | 0.89872 | 284* |
| Participant's Age at first regular cycles | 13.0622 | 1.10221 | 284* |
| Participant's Height (cm) | 155.6267 | 7.89824 | 367 |
| Participant's Weight (Kg) | 59.6056 | 13.19651 | 367 |
| Participant's Body Mass Index (BMI)<br>(Kg/cm<sup>2</sup>) | 23.5869 | 4.81608 | 367 |

\* 83 Participants did not recall this information.

**Figure 1.**
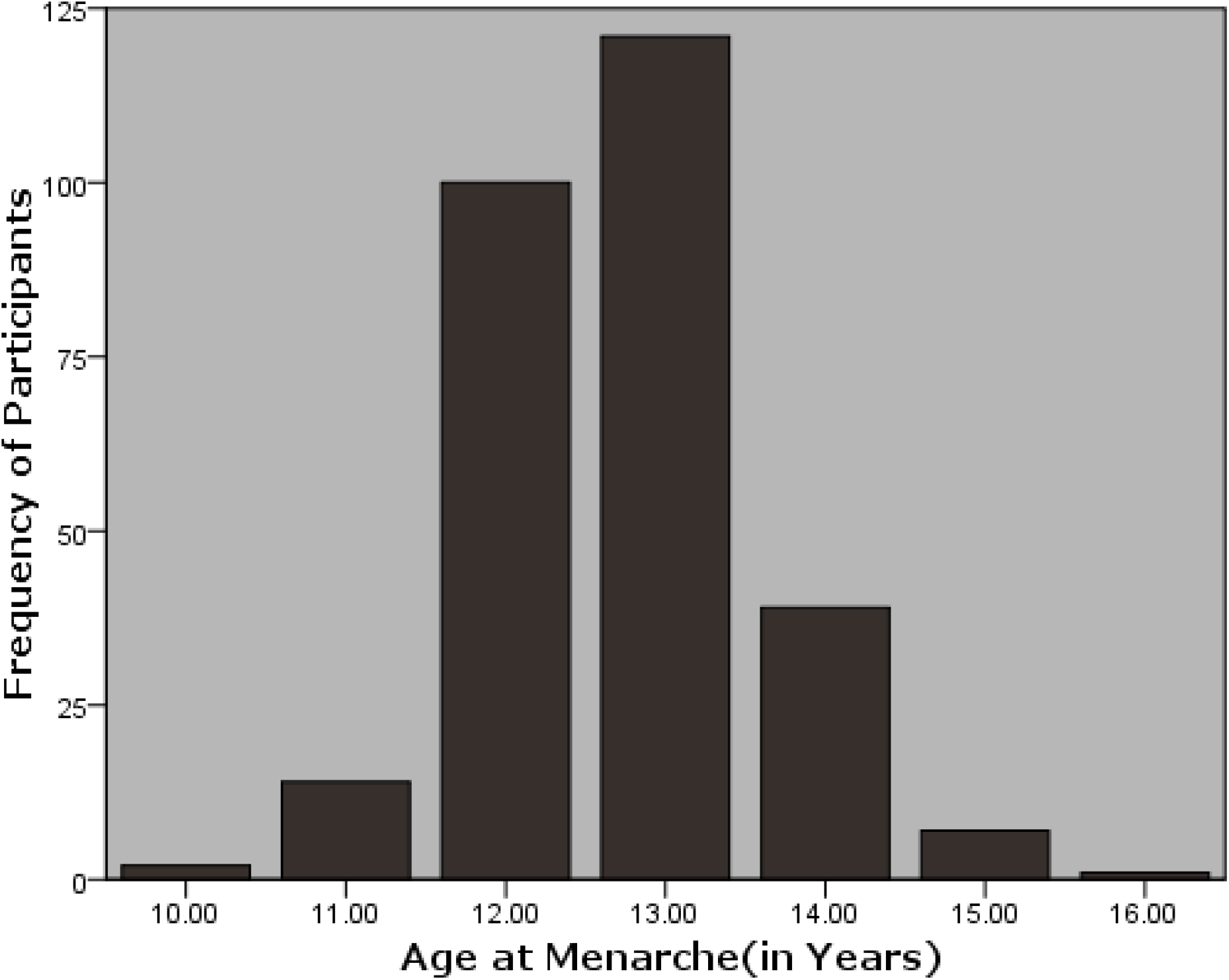
Age at menarche distribution among participants in years.

**Figure 2.**
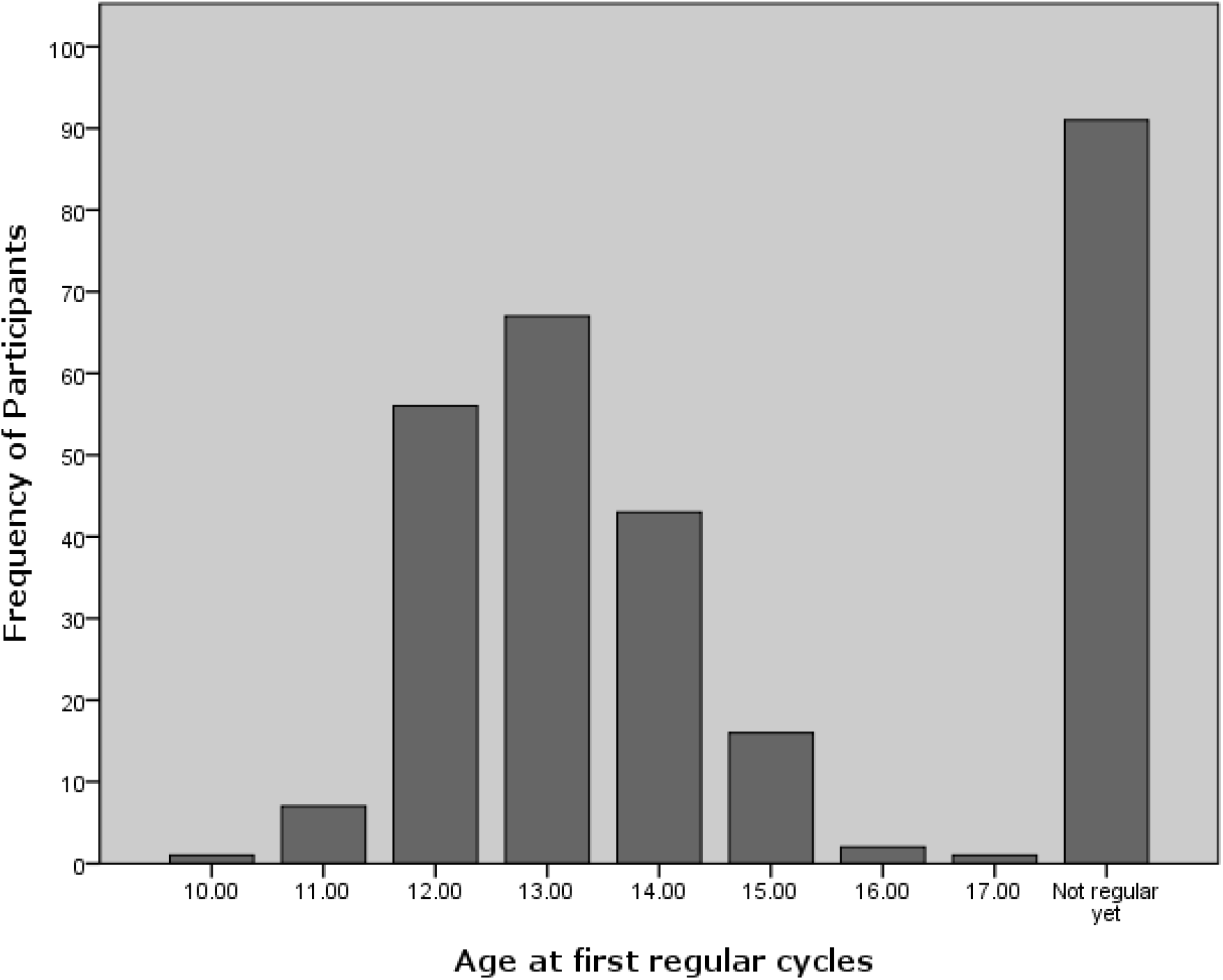
Participants Age at First regular Cycles

There seems to be a weak negative correlation between the age at menarche and weight in our study population. The same applies for the correlation between the SES and age at menarche. Nevertheless, there is a weak positive correlation between height and age at menarche.

## Discussion

In this study, the mean age at menarche was found to be 12.73 ± 0.89 years. The study by Attallah showed that it was 13.35 ± 0.14 years in the high income girls, 13.85 ± 0.15 years in the middle income, and 14.06 ± 0.18 years in the low income girls, in 1980.^[4]^ An implication of this is the possibility that, there is a decrease in the age of menarche and reproductive maturity for girls in the Sudan.^[5]^ This finding is in agreement with other results worldwide ^[6]^, a study in Spain showed that it decreased from 13.72 years to 12.83 years, between 1925 and 1962.^[7]^ In the Netherlands it declined from 13.66 years in 1955 to 13.15 in 1997.^[8]^ Similar studies in Mexico, China, and Indonesia, showed the same decrease in the menarcheal age.^(9–11)^

In our study the girls started to menstruate regularly at a mean age of 13.06 ± 1.10 years, which is less than a year after the first menstrual cycle. This contradicts the belief that nearly 1 to 2 years are needed for a girl to have regular cycles after menarche as it was linked to ovulation. ^[[9]^ However, there has been little discussion about the age at which girls tend to have regular cycles. One of these studies is a study done by Hosokawa et al, where they have demonstrated that 11.9% of their study women had regular cycles 1 year after their menarche.^[10]^ Moreover, researchers at the E3N-EPIC study found that about 31.6 % of their participants had regular cycles 1 year after menarche. In the first study, the participants were born in the 1980s and in the second, they were born between 1925 and 1930. ^[11]^ We asked a small size of the girls (not statistically significant) about their mother’s age at menarche and it showed the same age as the girls, which indicate that there is a genetic base for this finding, which needs more studies to clarify the link.

A strong relationship between weight and age at menarche have been reported in the literature.^[12, 13]^ Moreover, the increase in weight have been described as one of the factors that determine earlier menarche in females, including African females. However our study has found a weak reversible relation between them. Albeit there are few data and reports in African countries to draw a strong conclusion about this relationship. Although in recent years the amount of research regarding the matter are in the rise.^[2]^ On the other hand, this study has found a weak positive relationship between age at menarche and height. This variable drew the attention of health professions researchers, and its relation with menarche in general as well as with age at menarche. They agree with us on the positive correlation with age at menarche.^[14]^ But a national study from Colombia showed it to be inversely related to menarche, which is no popular finding among researchers who studied the relationship.^[15]^

The link between SES and age at menarche is substantial, as our findings suggest that it is an inverse and weak bond. Mohammed ElSheikh and Ammar Mohammed Ali Mohammed has recognized the same in northern Sudan, although they linked the age at menarche directly to the father and mother level of education and then separately to the economic status of the family.^[16]^ Similarly in a large cohort of immigrants in Europe SES was one of the major determinants of menarcheal status.^[17]^ In contrast to earlier findings, however, no significant relation was detected between SES and age at menarche in a collection of different ethnic groups by some researchers in USA.^[18]^

In conclusion, our study has determined the age of menarche in Sudanese girls albeit the size of our sample is small. It also made it obvious that there is a correlation, although it is weak, between age at menarche and SES in addition to weight and height. Further studies are needed for this important topic especially with the social, economic and cultural changes taking place in the Sudan and worldwide.

## Data Availability

All data produced in the present study are available upon reasonable request to the authors

